# *RV IsoMax:* Development of a web-based single-beat analysis tool for the determination of RV-PA coupling

**DOI:** 10.1101/2025.07.21.25331587

**Authors:** Shreya Agarwal, Alexandra Miller, Steven Hsu, Farbod N. Rahaghi, Scott Visovatti, Rebecca R Vanderpool

**Author notes:** **Corresponding Author:** Rebecca R. Vanderpool, PhD, Assistant Professor, Division of Cardiovascular Medicine, The Ohio State University, Davis Heart and Lung Research Institute, Suite 611b, 473 W. 12th Ave, Columbus, OH 43210, Twitter: @rrvdpool, Bluesky: @rrvdpool.bsky.social.

## Abstract

**Background:** Right ventricular (RV) function significantly associates with mortality in pulmonary hypertension. Single-beat methods allow for clinical estimates of max isovolumetric pressure (Pmax) but have not been widely adopted due to inter-user and inter-method variability and center specific implementation of methods. The study aim was to develop a web-based analysis single-beat analysis program that optimizes single-beat Pmax estimates while reducing inter-user variability.

**Methods:** Participants with a right heart catheterization were retrospectively identified from The Ohio State University Pulmonary Hypertension Registry. Right ventricular waveforms were digitized. The single-beat analysis program, *RV IsoMax*, was developed in R/RShiny to semi-automatically estimate the maximum isovolumetric pressure (Pmax). *RV IsoMax* results were compared to the popular 2^nd^ derivative single-beat method. Interclass correlation coefficients were calculated for interobserver and inter-method variability in Pmax estimates.

**Results:** The *RV IsoMax* method was tested across the spectrum of PH in participants with PH (n=45) and without PH (n=10). For the *RV IsoMax* method, there was no significant bias in Pmax estimates between observers (intraclass correlation coefficient: 0.96). In an inter-method comparison, the 2^nd^ derivative method significantly underestimated Pmax compared to *RV IsoMax* with a bias of 24 mmHg (intraclass correlation coefficient to 0.69). The variability between methods persists into end-systolic elastance (Ees) and RV-PA coupling (Ees/Ea).

**Conclusions:** The interactive user interface of the *RV IsoMax* application allows the user to semi-automatically calculate and explore variability in Pmax estimates. Significant differences in Pmax estimates between single-beat methods indicate their cut-off values cannot be used interchangeably.

**CLINICAL PERSPECTIVE:** *What is new?:*

- *RV IsoMax* is the first publicly available web-based tool for the single-beat analysis of RV pressure waveforms available at: https://vanderpoolrr.shinyapps.io/RV_IsoMax/.
- RV IsoMax is a semi-automatic algorithm that has been implemented with an easy-to-use graphical user interface (GUI) that allows the user to inspect the goodness of the Pmax estimations, adjust as needed and download the results as an excel file.
- In comparison to the 2^nd^ derivative single-beat analysis method, the RV IsoMax algorithm utilizes smaller fitting regions focused on early isovolumetric contraction and late isovolumetric relaxation that maximizes Pmax while allowing for reduced inter-user variability.

*What are the clinical implications?:*

- The web-based *RV IsoMax* app is a tool that can help facilitate the use of single-beat methods to determine RV-PA coupling in more clinical settings with standardized methods.

## INTRODUCTION

Right ventricular (RV) function is an important predictor of mortality in patients with pulmonary hypertension.^1,2^ Non-invasive assessments including RV ejection fraction and TAPSE/PASP are heavily used in clinical settings to assess right ventricular function but they global measures of RV function or surrogates of ventricular vascular coupling.^3,4^ Multi-beat pressure-volume (PV) loops are considered the gold standard method for assessing ventricular function with measures of ventricular contractility with end-systolic elastance (Ees), a more complete measure of ventricular afterload with arterial elastance (Ea) and the ventricular-vascular coupling ratio of Ees/Ea as the load independent measures of ventricular function. Ideally, pressure and volume are measured simultaneously using an invasive pressure-volume conductance catheter in combination with pre-load alterations like the Valsalva maneuver or inferior vena cava (IVC) balloon.^5^ The complexity of the pressure-volume conductance catheters and associated costs have led investigators to develop clinical estimates and single-beat methods to assess PV loops and RV function using standard clinical fluid-filled catheters, right heart catheterization derived stroke volume and CMR/Echo derived ventricular volumes.^6–11^

The single-beat methods can be broken down into these general steps: a) identification of a single beat RV waveform, b) isolation of the isovolumetric contraction and isovolumetric relaxation regions, and c) choosing specific all or some of the isovolumetric regions to fit with a sinusoidal curve^7^ or a 4th order Weibull distribution^12^. Beat to beat variability, inter-user variability and/or differences in single-beat algorithms influence the determination of Pmax. Multiple methods have been proposed for how to isolate the isovolumetric contraction and isovolumetric relaxation regions including the 1^st^ derivative method (dP/dt)^7^, the 2^nd^ derivative method (d^2^P/dt^2^)^8^ and the square of the 2^nd^ derivative method^12,13^.

Sunagawa et al. initially proposed and developed the simplified single-beat approach for estimating Pmax using the 1^st^ derivative method^14,15^ and Brimioulle S et al. then validated the method in anesthetized dogs.^7^ In the 1^st^ derivative method, the fittint regions are defined by max or min dP/dt and 10% of the max or dP/dt. Bellofiore A. et al proposed the 2^nd^ derivative method to reduce the inter-user variability.^8^ The main difference in the 2^nd^ derivative method is that the fitting regions are defined by the max and min 2^nd^ derivatives surrounding max and min dP/dt points in contraction and relaxation, respectively. Both methods have to further investigate RV function and RV-PA coupling in patients with pulmonary hypertension.^6,8,9,11,16^ Recent single-beat method development has focused on the 2^nd^ derivative method because of good interobserver agreement (ICC: 0.98). However, a significant limitation of 2^nd^ derivative method is it will consistently underestimate Pmax.^8^ Where the 1^st^ derivative method was not widely adopted due to difficulties with implementation and consistency in estimating Pmax. We have developed a web-based semi-automated method, *RV IsoMax*, to estimate Pmax that incorporates landmarks from the 1^st^ and 2^nd^ derivative methods to identify early isovolumetric contraction and late isovolumetric relaxation regions. We hypothesize that the *RV IsoMax* method will improve interobserver variability compared to earlier 1^st^ derivative methods while maximizing Pmax estimations compared to the 2^nd^ derivative method.

## METHODS

### Study population

A retrospective study population was used to study the applicability of the web-based analysis program on screen captures of RV pressure waveform data. Participants were identified at the Ohio State University (OSU) Medical Center between the years 2010-August 2021 with cardiac magnetic resonance imaging and right heart catheterization data (N = 61). The study was approved by The Ohio State University’s Institutional Review Board and informed consent was waived (IRB# 2021H0394).

### Digital Conversion of RV Pressure Screen Captures

Screen captures of RV pressure waveforms were collected from the right heart catheterization report during retrospective chart review (**Figure S1**). RV pressure screen captures were digitized to extract numerical time and pressure data from the images. In the digitization process, markers were placed to capture the RV pressure-time curve with additional markers placed around local minima, local maxima and inflection points. To calibrate time and pressure, pixels were scaled in the y-direction based on pressure scale and in the x-direction based on the time scale. Data was exported and saved as a text file for further analysis.

### Web-based program development

In the development of the web-based single-beat analysis program, we focused on improving the automation and reproducibility of the determination of Pmax. Program design considerations included: (1) the need for a graphical user interface to review the quality of the sinusoidal fit and make minor adjustments; (2) the capability to segment the RV pressure waveform and calculate Pmax automatically/semi-automatically; (3) inclusion of performance metrics and error checking (4) the ability to save analysis and figures; (5) conducting the analysis locally to maintain patient data security; (6) enabling the program to be platform-independent and function without the need for expensive licenses or large support files; and (7) provides added value to current practices (**Figure 1**). Based on previous success in deploying a web-based analysis program^17^, R/RShiny was used to develop the web-based single-beat RV function analysis program. The format for the input file is an Excel workbook with time (in milliseconds) in the first column and pressure (in mmHg) in the second column (**Figure S1**).

**Figure 1.**
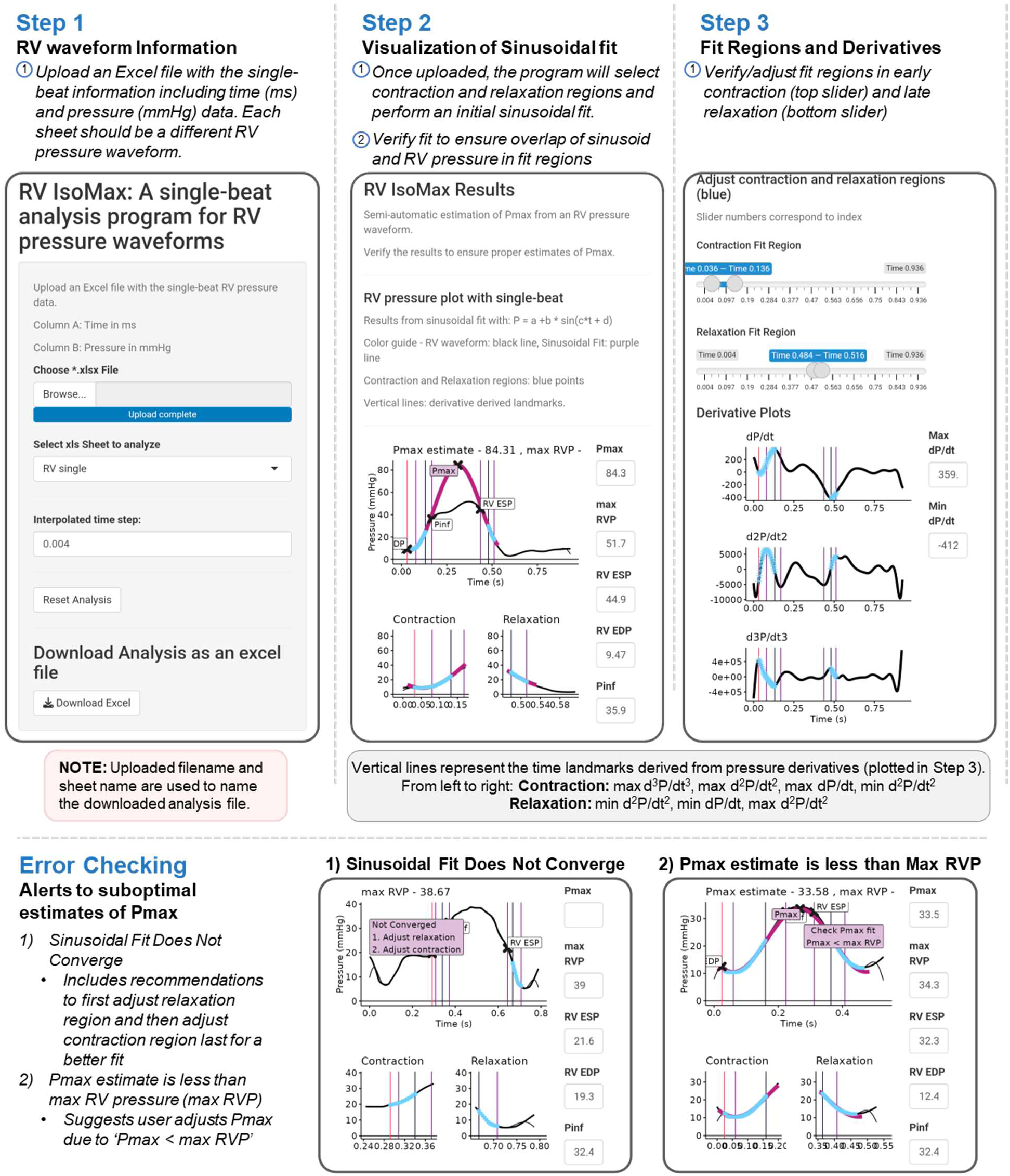
Screenshots of the *RV IsoMax* analysis program on a mobile device. The *RV IsoMax* analysis program allows the user to upload a single RV pressure waveform and then perform single-beat analysis to determine the max isovolumetric pressure (Pmax). The steps to use the application include: **Step 1:** The user uploads an excel file with the digital time (in milliseconds, ms) and pressure (in mmHg) in the first and second column respectively. The default is to perform single-beat analysis on the first sheet in the excel file but the user can select a different sheet if desired. This initial section of the application also allows the user to adjust the time step, to reset the analysis to the initial selection and to download the completed analysis as an Excel file. **Step 2:** The initial estimates of Pmax from the *RV IsoMax* application are displayed including the Pmax estimate, RV pressure landmarks (RV end-diastolic pressure, pressure at inflection, and RV end-systolic pressure) and the Sine curve (Purple line) overlaid on the RV pressure waveform (Black line). The early contraction and late relaxation regions are highlighted to allow the user to evaluate the fit regions (blue points) and the fit of the Sine curve to the original pressure. **Step 3:** The user can adjust the ‘Contraction Fit Region’ or the ‘Relaxation Fit Region’ if the initial fit is not optimal. The 1^st^, 2^nd^ and 3^rd^ pressure derivatives are also displayed to give the user additional guides when selecting points. Once the user is satisfied with the results, they can return to the options in Step 1 to download the analysis including images as an excel file. **Error Checking:** Two common errors that cause suboptimal estimates of Pmax have been identified and alerts have been programmed into the program. In the first instance, the sinusoidal fit does not converge using the automatically detected fit regions. In this case, the program will display the RV waveform, the fit regions and an alert that Pmax did not converge. The second common error is that the Pmax estimate is less than max RV pressure. In this case, the program displays the alert to ‘Check Pmax fit, Pmax < max RVP’.

### RV IsoMax Algorithm: a semi-automated method for identification of isovolumetric contraction/relaxation

The *RV IsoMax* algorithm is the primary algorithm used in the web-based single-beat analysis program/app. The *RV IsoMax* algorithm builds off previous methods to segment the RV pressure waveform with the addition of the third derivative to identify RV end-diastolic pressure.^18^ To reduce some of the noise in the derivatives, the pressure signal is first run through a Butterworth filter before calculating the first, second and third pressure derivative (**Figure 2A**). The start of contraction and end-diastolic pressure (pt 1) was identified using the third derivative (d3P/dt3) as the first maximum of d3P/dt3 that precedes max dP/dt. The maximum and minimum of the first derivative are identified to mark the end of contraction (pt 2: max dP/dt) and start of relaxation (pt 3: min dP/dt). From the second derivative (d2P/dt2), the end of relaxation (pt 4) was identified as the first maximum of d2P/dt2 that follows min dP/dt. Additional points identified include pressure at first inflection (*Pinf:* first minimum of d2P/dt2 that follows max dP/dt) and RV end-systolic pressure (*ESP:* minimum of d2P/dt2 that precedes min dP/dt).

**Figure 2.**
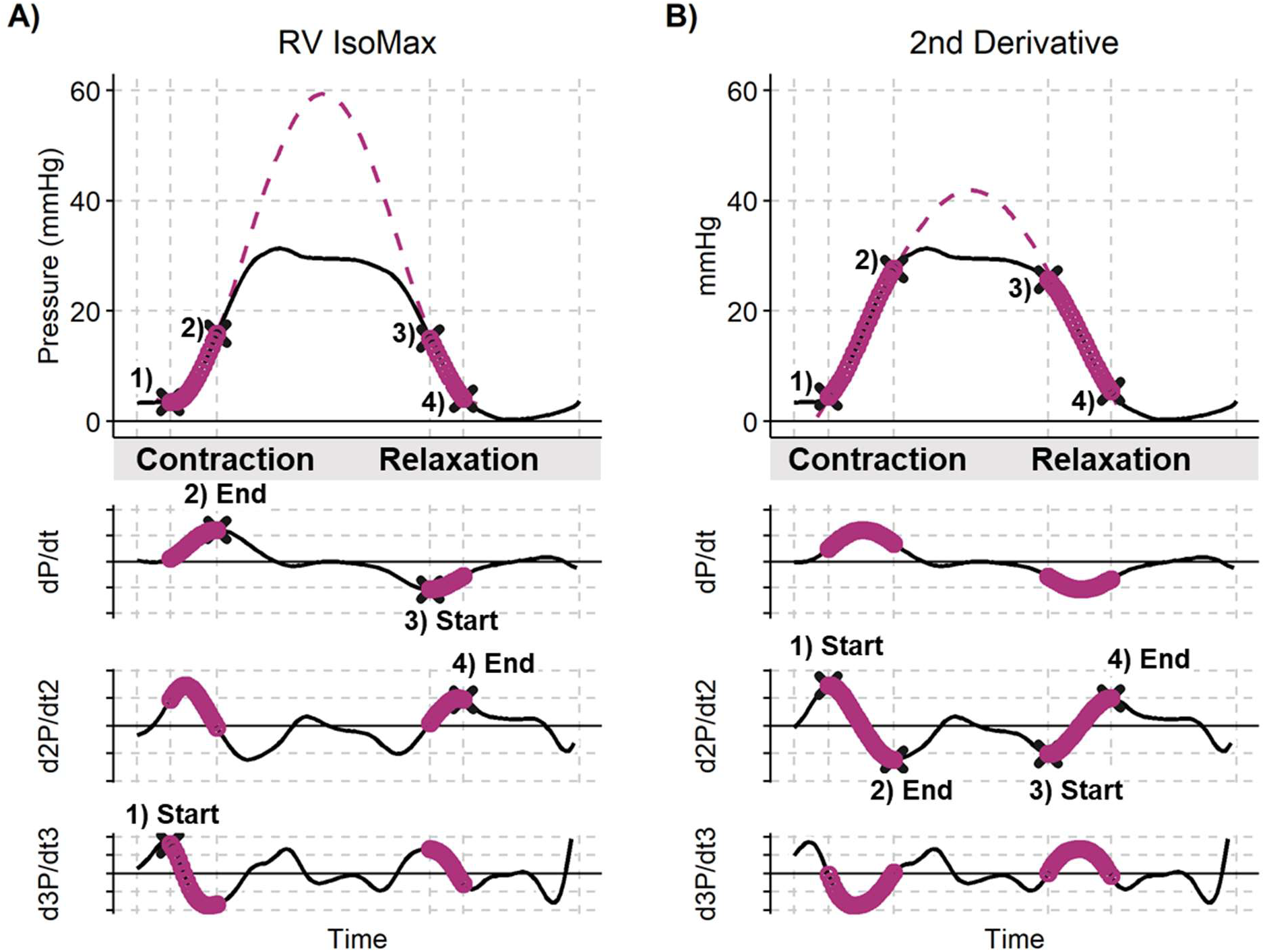
Comparison of *RV IsoMax* and 2^nd^ Derivative single-beat methods for the estimation of max isovolumetric pressure, Pmax. **A)** In the *RV IsoMax* method, the start of the contraction region (pt 1) is defined by the max of the 3^rd^ derivative before max dP/dt. The end of the contraction region (pt 2) is defined by max dP/dt. The start of the relaxation region (pt 3) is defined as min dP/dt and the end of the relaxation region (pt 4) is defined by the max of the 2^nd^ derivative after min dP/dt. **B)** In the 2^nd^ derivative method, all regions are defined using 2^nd^ derivative landmarks. The contraction region (pts 1-2) is defined using the region between max and min of the 2^nd^ derivative that surround max dP/dt. The relaxation region (pts 3-4) is defined using the region between min and max of the 2^nd^ derivative that surround min dP/dt.

### Second Derivative Method

Pmax estimates from the *RV IsoMax Algorithm* were compared to those derived from the second derivative method.^8,11,12^ In the second derivative method, the contraction and relaxation regions were all derived from the second derivative. Briefly, the contraction region was defined as starting at the first maximum of d2P/dt2 that precedes max dP/dt and ending at the first minimum of d2P/dt2 that follows max dP/dt (**Figure 2B**, pts 1 and 2). The relaxation region was defined as starting at the first minimum of d2P/dt2 that precedes min dP/dt and ending at the first maximum of d2P/dt2 that follows min dP/dt (**Figure 2B**, pts 3 and 4). Alternatively, the square of the second derivative can be taken to produce four upright peaks to define the pressure segments used for Pmax predictions.^11^

### Hemodynamic and Single-Beat RV function variables

Hemodynamic measures of RV function were determined including Ees, Ea, and Ees/Ea, (**Table S1**). A pressure-only based measurement of RV ejection fraction was also calculated as RVEF_est_ = 1 – (ESP/Pmax).^12^ A major assumption of the pressure-only estimate of RVEF is that the unstressed volume of the RV (V_0_) is zero. However, it has been previously shown that V_0_ significantly increases with pulmonary hypertension.^6^ The pressure-only estimate of RV ejection fraction was compared to the CMR derived RVEF.

### Statistical Analysis

Results are expressed as median (interquartile range) or mean ± SD, where appropriate. Comparisons between groups were made using a t-test or Mann–Whitney test, where appropriate. P-values <0.05 were considered significant. Bland–Altman and intraclass correlation coefficient analyses were used to compare single-beat results between methods (*RV IsoMax* and 2^nd^ Derivative) and to determine the inter-observer variability. For survival analysis, follow-up time was calculated from the date of the RHC to the date of death or censoring. Cox proportional hazard regression was used to compute hazard ratios (HR) and 95% confidence interval (CI) for death. Receiver Operator curves were used to identify optimal cut-offs for Ees/Ea ratios and Kaplan-Meier curves were generated. The statistical analyses were performed using R programming language (r-project.org, version 4.2.3) and RStudio (version 2024.09.0+375).

## RESULTS

Sixty-one participants were identified with CMR imaging, invasive RHC and RV waveform data in the OSU CMR PH registry. Ten participants with normal pulmonary artery pressure were identified (No PH, age: 53±17 years, sex: 60% female). Majority of participants with pulmonary hypertension were classified as having WSPH group 1 (n = 29, age: 54±15 years, sex: 76% female, **Table 1**).

**Table 1.** Patient characteristics. Categorical variables as n (%) and continuous variables and mean ± standard deviation.

|  | Overall |
| --- | --- |
| Women | 41 (67.2) |
| Age, y | 55 ± 15 |
| WSPH Group |  |
| No PH | 10 (16.4) |
| WSPH Group 1 | 29 (47.5) |
| WSPH Group 2 | 16 (26.2) |
| WSPH Group 3 | 3 (4.9) |
| WSPH Group 4 | 2 (3.3) |
| WSPH Group 5 | 1 (1.6) |
| Deaths | 28 (45.9) |
| BSA, m<sup>2</sup> | 1.99 ± 0.31 |
| RA pressure, mmHg | 11 ± 7 |
| mPAP, mmHg | 40 ± 15 |
| PAWP, mmHg | 15 ± 9 |
| PVR, mmHg/L/min | 5.72 ± 4.43 |
| RV ejection Fraction, % | 41.4 ± 14.0 |
| RV end systolic pressure | 58 ± 25 |

**Table 2.** Comparison advantages and disadvantages of *RV IsoMax* and 2^nd^ Derivative methods for estimating Pmax.

|  | *RV IsoMax* Method | 2<sup>nd</sup> Derivative Method |
| --- | --- | --- |
| Advantages | • Includes only the early contraction and late relaxation regions maximizing Pmax estimates<br>• Minimal adjustments improve the fit when signals are noisy | • Only requires calculation of 2<sup>nd</sup> derivative<br>• Good convergence of Pmax estimations due to larger regions used in the non-linear fit. |
| Disadvantages | • Requires more data conditioning and can get complicated outside of scripting data analysis programs.<br>• Sensitive to noisy RV signals for detecting landmarks | • Pmax estimates significantly lower than *RV IsoMax* estimates<br>• No guidance for adjustment of points.<br>• Sensitive to noisy RV signals for detecting landmarks |

### RV IsoMax Single-Beat Web-based program/app to analyze RV pressure waveforms

For the *RV IsoMax* single-beat analysis program, we focused on development of it in R/RShiny based on previous design considerations for an interactive web-based tool.^17^ The current version of the *RV IsoMax* is designed to be usable on the desktop or laptop computer and on mobile devices (**Figure 1**). Additional information about the *RV IsoMax* analysis program is available on our GitHub repository (https://github.com/vanderpoolrr/RV_IsoMax). The end-user begins with uploading an excel file that contains the single-beat RV pressure waveform with time in the first column (in milliseconds, ms) and pressure in the second column (in mmHg). The program will then identify the number of sheets present in the uploaded file and will begin analysis on the first sheet. Behind the scenes, the program takes the 1^st^, 2^nd^ and 3^rd^ derivative of the RV pressure waveform and identify the early contraction and late relaxation regions on the waveform and display the results in steps 2 and 3 (**Figure 1**). The sinusoidal curve fit will then be performed using those contraction and relaxation regions (blue symbols) and the sinusoidal results (purple curve) and Pmax are displayed in relation to the original RV waveform (black curve). In Step 2, the user can verify the single-beat results are good and can adjust the fit in step 3 if needed. If the default regions are not suitable for the sinusoidal curve fit, the user can adjust the Contraction and Relaxation Fit Regions using the sliders (**Step 3**). The vertical lines in the figures represent the identified derivative landmarks including RV end-diastolic pressure, inflection pressure (Pinf), and RV end-systolic pressure (RV ESP). Once the user has come to a suitable fit, the results including the figures, RV waveform, derivative and fit regions can be downloaded as an excel file (**Figure S2**).

Error checking and warning messages have also been integrated to guide the user to the best fit (**Figure 1**, Error Checking). A common error is that the fit of the sinusoidal curve does not converge to a solution with the automatically identified regions. In this case, the RV waveform and the identified regions will be displayed with a warning that the fit did not converge and suggest the user first adjusts the relaxation region followed by the contraction region. The second common error is the when the estimate of Pmax is less than the maximum pressure in the original RV waveform. This should prompt the user to adjust the fit regions to obtain a better fit.

### Stability of Pmax estimations: Comparison of RV IsoMax to 2^nd^ Derivative Methods

To assess the stability of the Pmax estimates, inter-user variability between two observers (RRV and SA) using the *RV IsoMax* analysis program were determined (**Figure 3A**). There were no significant differences in the average Pmax with minimal bias (1.3 mmHg) and small limits of agreement of -27.1 to 29.8 mmHg between observers (**Figure 3A**). The Intraclass correlation coefficient (ICC) showed excellent reliability between observers with an ICC of 0.965 (95% confidence interval: 0.942-0.979). When the *RV IsoMax* and 2^nd^ derivative methods were compared there were significant inter-method variability (**Figure 3B**). Pmax estimates were significantly lower for the 2^nd^ derivative method (P< 0.001) with significant biases and large limits of agreement (bias: 24 mmHg, limits of agreement: -45 to 93 mmHg). There was moderate reliability between methods with an ICC of 0.687 (95%-Confidence Interval: 0.345-0.839). The variability persists into the end-systolic elastance (Ees) and RV-PA coupling (Ees/Ea) calculations (**Figure 3C and D**). The 2^nd^ derivative method underestimates Ees (bias: 0.33 mmHg/ml and limits of agreement: -0.91 to 1.57 mmHg/ml) and Ees/Ea compared to *RV IsoMax* (bias: 0.48 and limits of agreement: -0.73 to 1.69).

**Figure 3.**
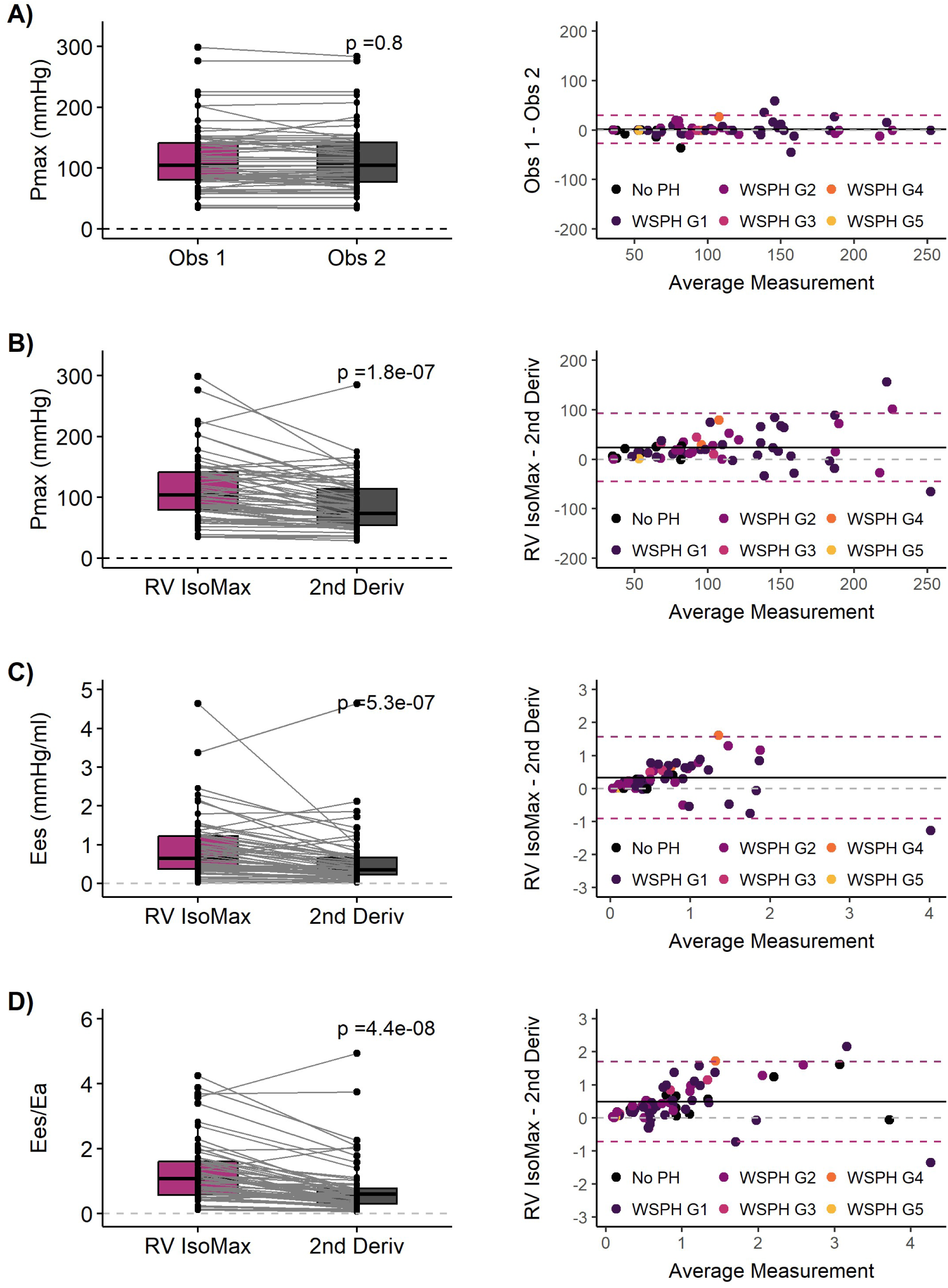
Validation and comparison of the single-beat methods for determining Pmax. **A)** Assessment of interobserver variability based on the average Pmax (left) and Bland Altman analysis (right). Colors in the Bland-Altman plot indicate World Symposium of Pulmonary Hypertension (WSPH) group assignment. **B)** Comparison of Pmax estimates using the *RV IsoMax* and 2^nd^ derivative method (2^nd^ Deriv) based on average between methods(left) and Bland Altman analysis (right). Calculations of end-systolic elastance **(C)** and RV-PA coupling ratio Ees/Ea **(D)** were also compared.

### Challenging Waveforms and Suggested Approaches

Despite automatic segmentation and detection of landmarks some waveforms are difficult to fit for a variety of reasons. We have identified three examples of difficult fits that had the largest difference in the Pmax estimates between observers and represent typical challenging cases. In the first example, there were multiple local positive dP/dt peaks in the isovolumetric contraction region (**Figure 4A**). Multiple local positive peaks in the middle of the isovolumetric contraction region can confuse the algorithm. Normally, max dP/dt will be in the early contraction region right after RV end-diastolic pressure. In this example, the largest peak is the second positive dP/dt peak that results in an over-estimation of RV end-diastolic pressure and using the second half of the contraction period. In order to obtain a Pmax estimation, the user should adjust the contraction region to span the more accurate RV end-diastolic pressure and the first dP/dt peak (**Figure 4B**). Another difficult example is when there is a significant increase in RV end-diastolic pressure resulting in strongly asymmetric waveforms (**Figure 4C**). Asymmetric waveforms are cases when the 2^nd^ derivative method is better for determining Pmax because it includes a larger fitting range^8^ (**Figure 4D**). Visual inspection of the fitting curve with manual adjustments is also possible in these cases but can lead to more subjectivity in the corrections and more variability. In the final example, there was an initial Pmax estimation with convergence of the fit but it seems to underestimate Pmax. Visual inspection of the fit at the start of contraction shows the sinusoidal curve crossing/underestimating diastolic pressure (**Figure 4E**).

**Figure 4.**
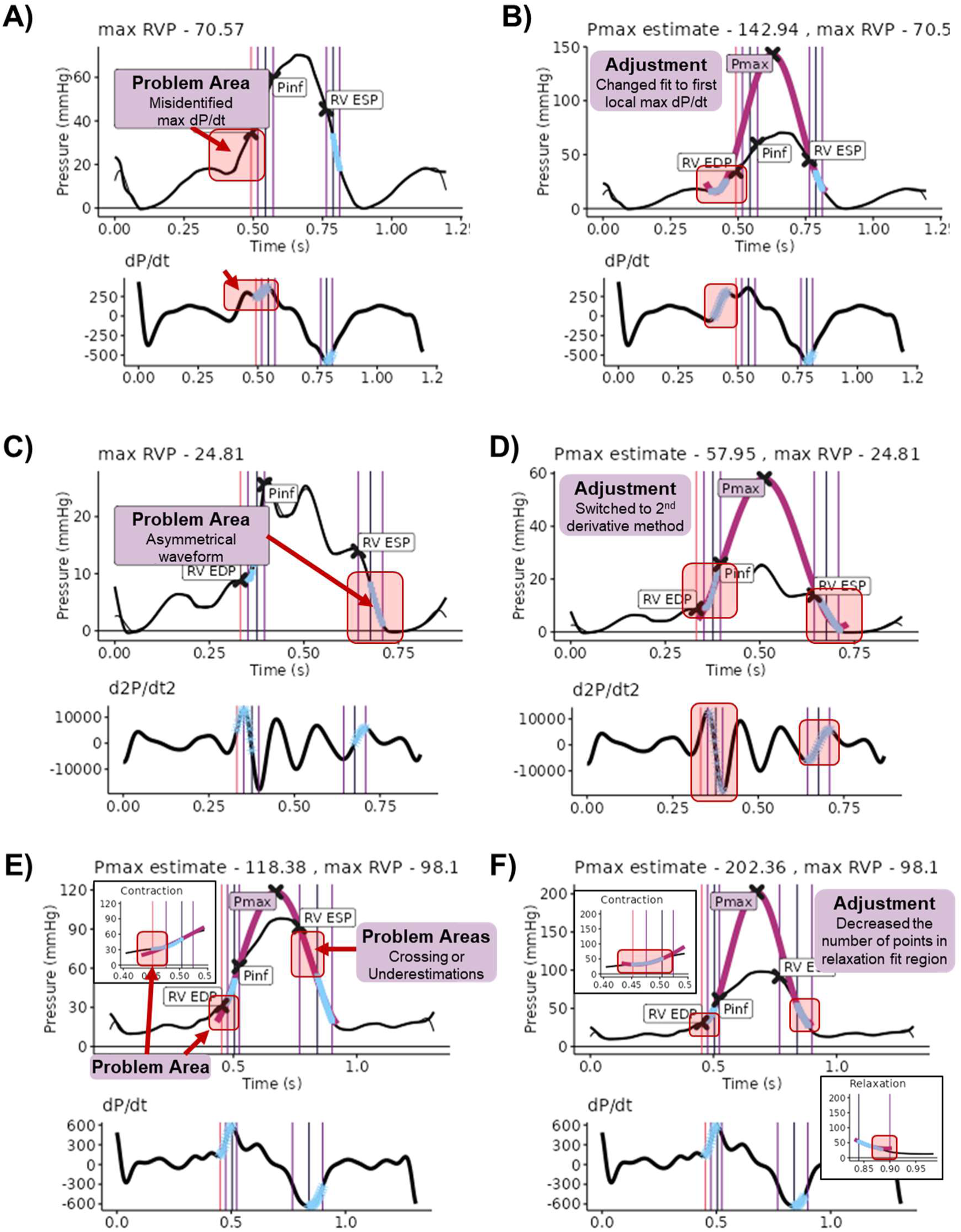
Identification of problem areas and resulting adjustments in difficult to fit RV pressure waveforms. **A)** In the first example, the program misidentified the max dP/dt (second local peak) as the peak dP/dt in the early contraction region. This resulted in an asymmetric waveform/fit and no convergence of the Sine curve. **B)** The solution is to adjust the contraction fit region to the first local dP/dt peak and that results in a good Pmax estimation. **C)** In the second example, the waveform is asymmetric with a high RV end-diastolic pressure. **D)** The proposed adjustment in this case is to switch to the 2^nd^ derivative method to incorporate more of the contraction and relaxation regions in the fit. **E)** In the third example, there is a Pmax estimate but the overlay of the Sine curve (purple curve) on the original RV waveform (black curve) shows the Sine curve crosses and underestimates RV pressures at the end of diastole. Additionally, the Sine curve is right on top of the original RV waveform during early relaxation. **F)** The solution is to adjust the end of the relaxation fit region. Problem areas are highlighted using red arrows and red boxes. Adjustments are highlighted using a red box.

Additionally, the sinusoidal curve is right on top of the RV pressure waveform before min dP/dt. They type of fit is likely due to a small amount of asymmetry in the waveform (**Figure 4E**). In this case, visual inspection of the fit with minor adjustments at the end of the relaxation region result in a much higher estimate of Pmax (202 vs 118 mmHg). Additional work is needed to refine the recommended actions in these cases to get the best Pmax estimates while also minimizing variability.

### Association of Ees/Ea with mortality and changes in RV-PA coupling at follow-up

As continuous variables, Ees/Ea did not associate with death in this cohort (*RV IsoMax* Ees/Ea: HR: 0.55 and 95% CI: 0.22-1.37 and 2^nd^ derivative Ees/Ea: HR: 0.52 and 95% CI: 0.14-1.94). Based on ROC analysis, the optimal cut-off for *RV IsoMax* was 1.0 and for the 2^nd^ derivative method was 0.3. The Kaplan Meier analyses, were not significant with p-values of 0.064 and 0.095 (**Figure S3**). The change in Pmax from an index RHC to 6-year follow-up was assessed one subject using both methods (**Figure 5**). Using the *RV IsoMax* method, Pmax decreased from 206 to 172 mmHg with an improvement in Ees/Ea from 1.22 to 1.97. For the same individual using the 2^nd^ derivative method, Pmax decreased from 138 to 118 mmHg with an improvement in Ees/Ea from 0.48 to 1.03. Suggesting both methods would capture the improvements of RV function that is reflected in the increase in stroke volume from 40 to 60 ml over the same time period. However, the absolute values and magnitude of change are different between the two methods.

**Figure 5.**
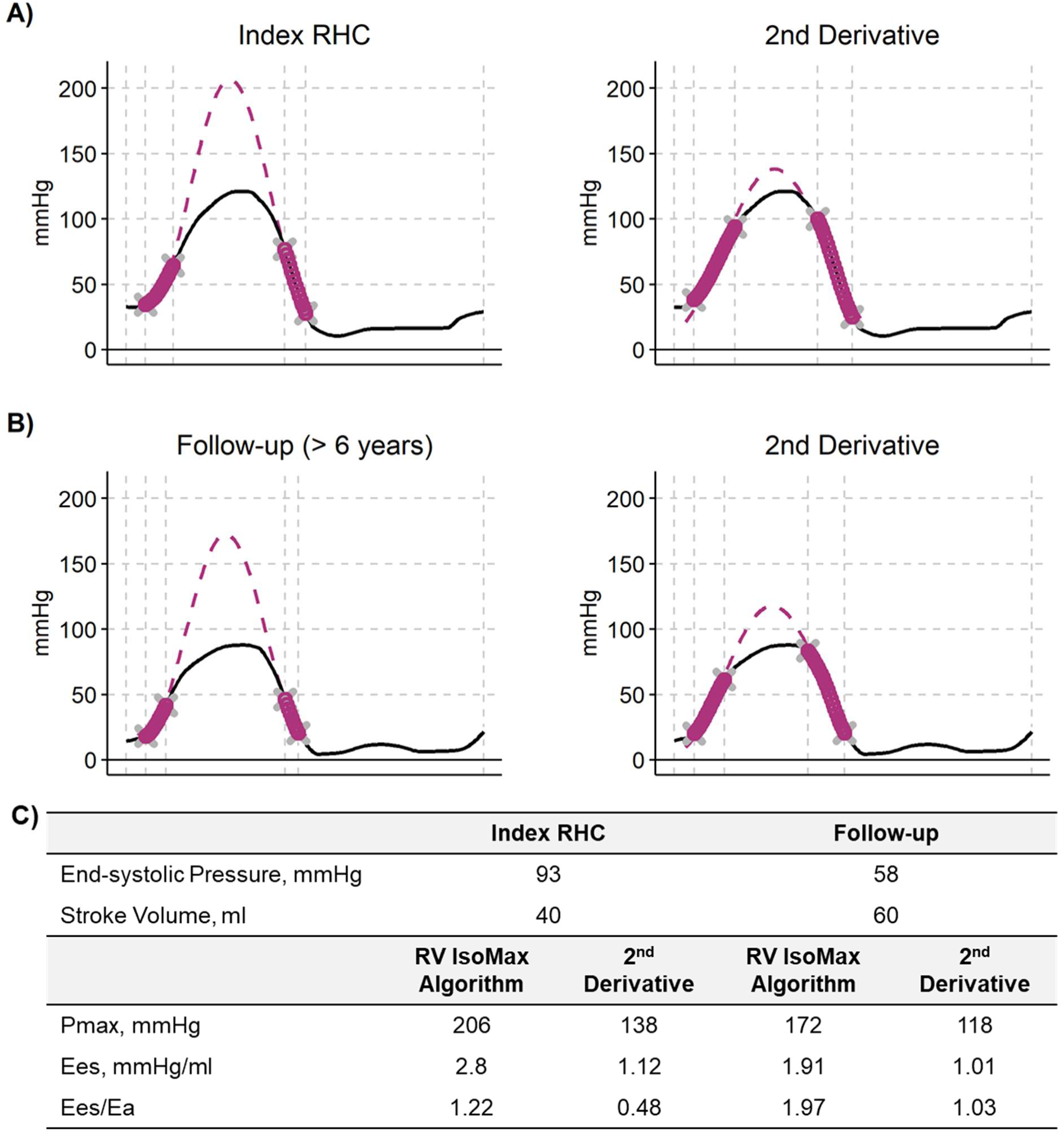
Comparison of Pmax single-beat estimates at follow-up. At both the index **(A)** and follow-up **(B)** right heart catheterization, *RV IsoMax* method had higher estimates of Pmax **(C)**. Estimates of Pmax significantly impact RV contractility (Ees) and RV-PA coupling (Ees/Ea) suggesting methods and cut-offs are not interchangeable.

## DISCUSSION

In this study, we developed *RV IsoMax*, a web-based analysis tool, to analyze digital RV pressure waveforms for the estimation of Pmax and RV-PA coupling in patients with pulmonary hypertension. This implementation of the single-beat method should help with the standardization of the single-beat assessments of RV function in clinical setting. The web-based tool was successfully deployed to calculate Pmax with good agreement between different users. There were significant differences between the *RV IsoMax* and 2^nd^ derivative methods for estimating Pmax with the 2^nd^ derivative method estimating lower Pmax values.

### Comparison of Single-beat Methods

In the development of the *RV IsoMax* analysis tool, we have improved on the previous limitations of the 1^st^ derivative method by providing better guidance for defining the early contraction and late relaxation regions. There was excellent reliability between observers in the Pmax estimations (ICC: 0.965, CI 0.942-0.979) that were similar to previously reported reliability for the first derivative method (0.947, CI 0.892–0.970).^8^ While the 2^nd^ derivative method has excellent reliability with an ICC coefficient of 0.980 (CI 0.973–0.985), it significantly underestimates Pmax compared to the 1^st^ derivative method and *RV IsoMax* methods by 13% and 18% respectively (**Figure 3**).^8^ In the early validation of MRI-RHC derived RV pressure-volume loops, Kuehne T et al. used Brimioulle’s 1^st^ derivative single-beat methods to determine Pmax in 6 patients without PH and 6 patients with PH. ^7^ They found RV-PA coupling was decreased in PH patients (n = 6, 1.1±0.3) compared to controls (1.9±0.4; P<0.01).^16^ Using the *RV IsoMax* method of estimating Pmax, we found similar average Ees/Ea values for WSPH group 1 (Ees/Ea 1.27 ± 0.91) and patients without PH (Ees/Ea 1.8 ± 1.21) (**Table 3**). For the same subjects, 2^nd^ derivative derived Ees/Ea values were significantly lower for WSPH group 1 (0.84 ± 0.95) and patients without PH (1.26 ± 1.05).

**Table 3.** Single-beat results in patients without PH (No PH) and in World Symposium Pulmonary Hypertension (WSPH) groups 1 and 2 in the cohort. Patients with WSPH groups 3-5 were not included in the table due to small numbers. Continuous variables are presented as mean ± standard deviation. * P<0.05 vs No PH and † P< 0.05 vs WSPH 1

|  | No PH | WSPH 1 | WSPH 2 |
| --- | --- | --- | --- |
| N | 10 | 29 | 16 |
| RV EDP, RV end-diastolic pressure, mmHg | $7 \pm 3$ | $20 \pm 8^*$ | $18 \pm 8^*$ |
| P<sub>inf</sub>, Pressure at inflection, mmHg | $25 \pm 6$ | $56 \pm 19^*$ | $49 \pm 14^*$ |
| Max RVP, Max RV pressure, mmHg | $32 \pm 8$ | $76 \pm 25^*$ | $68 \pm 19^*$ |
| RV ESP, RV end-systolic pressure, mmHg | $24 \pm 9$ | $62 \pm 24^*$ | $59 \pm 18^*$ |
| ESP<sub>est</sub>, Estimated end-systolic pressure, mmHg | $20 \pm 6$ | $68 \pm 21^*$ | $62 \pm 21^*$ |
| Max dP/dt, mmHg/sec | $287 \pm 92$ | $537 \pm 235^*$ | $422 \pm 217$ |
| Min dP/dt, mmHg/sec | $-232 \pm 105$ | $-646 \pm 244^*$ | $-547 \pm 307^*$ |
| RVEF<sub>CMRI</sub>, CMR derived RV ejection fraction, % | $56 \pm 10$ | $38 \pm 12^*$ | $41 \pm 14^*$ |
| **RV IsoMax** |  |  |  |
| P<sub>max</sub>, Max isovolumetric pressure, mmHg | $61 \pm 20$ | $132 \pm 55^*$ | $117 \pm 60^*$ |
| E<sub>es</sub>, End systolic elastance, mmHg/ml | $0.44 \pm 0.23$ | $1.12 \pm 0.96$ | $0.77 \pm 0.72$ |
| E<sub>a</sub>, Arterial elastance, mmHg/ml | $0.28 \pm 0.10$ | $1.09 \pm 1.00^*$ | $0.75 \pm 0.36$ |
| E<sub>es</sub>/E<sub>a</sub>, RV-PA coupling ratio | $1.80 \pm 1.21$ | $1.27 \pm 0.91$ | $1.02 \pm 0.91$ |
| V<sub>0</sub>, unstressed RV volume, ml | $1 \pm 29$ | $38 \pm 81$ | $-74 \pm 292$ |
| RVEF<sub>Pressure</sub>, Pressure derived RV ejection fraction, % | $59 \pm 15$ | $50 \pm 16$ | $43 \pm 20$ |
| **2<sup>nd</sup> Derivative** |  |  |  |
| P<sub>max</sub>, Max isovolumetric pressure, mmHg | $48 \pm 12$ | $107 \pm 49^*$ | $92 \pm 39^*$ |
| E<sub>es</sub>, End systolic elastance, mmHg/ml | $0.28 \pm 0.15$ | $0.79 \pm 0.92$ | $0.44 \pm 0.40$ |
| E<sub>a</sub>, Arterial elastance, mmHg/ml | $0.28 \pm 0.10$ | $1.09 \pm 1.00$ | $0.75 \pm 0.36$ |
| E<sub>es</sub>/E<sub>a</sub>, RV-PA coupling ratio | $1.26 \pm 1.05$ | $0.84 \pm 0.95$ | $0.56 \pm 0.47$ |
| V<sub>0</sub>, unstressed RV volume, ml | $-53 \pm 86$ | $-44 \pm 152$ | $-266 \pm 465^\dagger$ |
| RVEF<sub>Pressure</sub>, Pressure derived RV ejection fraction, % | $49 \pm 18$ | $38 \pm 17$ | $31 \pm 17$ |

Single-beat methods show good agreement with the gold-standard multi-beat pressure volume analysis when Ees/Ea is less than 1 and RVEF is less than 40%.^19,20^ In one study, Richter MJ et al. found good agreement between multi-beat and single-beat methods with an ICC of 0.95 and a 95% confidence interval of 0.89-98).^19^ In another study Hsu S. et al. did not find good agreement between methods across the full range of Ees/Ea and RV ejection fraction. They found the bias increased in participants with more normal RV function (Ees/Ea > 1 and RVEF>40%).^19^ These findings would suggest that the 2^nd^ derivative method would further underestimate Pmax and RV-PA coupling compared to multi-beat methods. Some of the variability in agreement between methods could be a result of different implementations of the 1^st^ derivative method at each center.

### Standardizing Single-beat Analysis

The *RV IsoMax* analysis program is an initial step in developing a single-beat analysis program that is accessible across centers. Single-beat methods have not been more widely used in clinical setting and management of patients with PH, is the lack of a robust, standardized implementation.^8^ Each center might use different single-beat methods (1^st^ derivative, 2^nd^ derivative and novel methods) in addition to different analysis programs (Matlab, SigmaPlot, R, Python, etc.). In comparison to left ventricular pressure waveforms, the RV pressure waveform significantly changes shape with increased afterload^21^ and PA occlusions^11,22^. The *RV IsoMax* program has been specifically designed and tested on RV pressure waveforms to overcome the typical challenges including shorter contraction and relaxation regions that can result in more unstable fits.^8^ Analysis of the second derivative allows for the identification of pressure landmarks and phases of the cardiac cycle for hemodynamic metrics of RV function including RV end-systolic pressure, diastolic function, filling and ejection times.^18^ A previous version of the program and algorithm was used to assess RV contractility and RV-PA coupling following left ventricular assist device implantation.^23^

### Estimating RV end-systolic pressure

Without the multi-beat pressure-volume loops, or measurements of time-resolved pressure and flow, it is difficult to determine end-systolic pressure or the unstressed volume of the right ventricle. Initial implementations of the single-beat method used mean PA pressure^6^ but later studies used conductance catheters in patients with PH to demonstrate that ESP increases with increased afterload to values that are closer to systolic PA pressure.^10^ Accordingly, the simple relationship of ESP = 1.65*mPAP-7.79 was proposed for when ESP measurements are not available. We can also estimate RV end-systolic pressure as the pressure at the start of the linear relaxation region when we use the 2^nd^ derivative of pressure to help identify RV landmarks.^11,18^

### Unstressed right ventricular volume, V_0_

Trip P. et al. demonstrated that the unstressed volume of the right ventricle (V_0_) significantly increases when there is significant RV remodeling as in patients with pulmonary arterial hypertension. ^6^ On average V_0_ was found to be 43 ml (range: -8 to 171) ml in patients with IPAH. Using *RV IsoMax* Pmax estimates, the average V_0_ in patients without pulmonary hypertension was 1± 29 ml and increased in WSPH group 1 (**Table 3**. V_0_: 38 ± 81ml). A potential reason for differences in V_0_ could stem from early studies using mean PA pressure as the estimate of RV ESP resulting in an overestimation of Ees and V_0_. Using the 2^nd^ derivative method, V0 was estimated as -53 ± 86 ml in patients without PH and did not significantly change in patients with WSPH group 1 (V_0_: -44 ± 152 ml).

### Sources of uncertainty in Pmax estimations

Uncertainties in Pmax estimations can arise at various stages of the single-beat analysis. During the single-beat identification stage, variability can occur due to the choice of representative beat or pre-processing of multiple beats to get an average representative beat. While not a focus of this manuscript, users should be careful they select a single beat that captures the variability across the respiratory cycle, especially if there are significant respiratory swing. Optimally, the use would include parts of the diastolic phase before and after the beat of interest. In the RV landmark and fit region identification stage, the *RV IsoMax* program has been designed to reduce the variability between users but errors can be introduced if the landmarks and subsequent fit regions are not accurately identified (**Figure 4A**). In the non-linear fit to estimate Pmax state, the fitting process and the chosen model could introduce errors if they do no adequately represent the true physiological behavior. In *RV IsoMax*, we focused on fitting a Sine curve to the RV pressure waveform^7,8^ but other groups have used a continuous probability distribution of a 4^th^ order Weibull distribution.^12,13^ When compared to the results using a Sine curve, the 4^th^ order Weibull distribution method gave similar results.

### Limitations

There are several limitations in the present study. The single-beat estimates of Ees/Ea were not compared to the gold-standard multi-beat derived Ees/Ea relationships. The focus of this manuscript was on the development of the web-based analysis program to standardize the implementation of single-beat methods. The current *RV IsoMax* program requires the user to have already identified and isolated a single RV pressure waveform to use. This was an intentional choice to not constrain the user to a single pressure source (XPER binary files, Mac-Lab export files or outputs from digitization programs). In this manuscript we used re-digitized RV waveforms that could have introduced more variability in results but they have previously been found to have similar characteristics to their original waveforms.^24^ RV pressure waveform data from patients with PH have been used in the development and testing of the current application. It is possible that other types of ventricular remodeling and cardiovascular disease could make the single-beat Pmax estimates more difficult like constrictive or restrictive cardiomyopathy that alter diastolic filling patterns.^25^

*Conclusions*.

The structured *RV IsoMax* algorithm and the web-based analysis program are a crucial step in transitioning single-beat Pmax estimations from specialized laboratory settings to the wider pulmonary hypertension (PH) community. Broader implementation of single-beat method should enhance our understanding of the intricate nature of right ventricular (RV) function and clinical relevance.

## Data Availability

All data produced in the present study are available upon reasonable request to the authors

https://github.com/vanderpoolrr/RV_IsoMax

## ABBREVIATIONS

dP/dt: 1^st^ derivative of pressure
d^2^P/dt^2^: 2^nd^ derivative of pressure
d^3^P/dt^3^: 3^rd^ derivative of pressure
Ea: pulmonary arterial elastance
Ees: End-systolic elastance
Ees/Ea: RV-PA coupling ratio
ESP: End-systolic pressure
Pinf: Pressure at the inflection point during systole
mPAP: mean Pulmonary Arterial Pressure
PAH: Pulmonary Arterial Hypertension
PAWP: Pulmonary Artery Wedge Pressure
PH: Pulmonary Hypertension
PAH: Pulmonary Arterial Hypertension
Pmax: Maximum isovolumetric pressure
PVR: Pulmonary Vascular Resistance
RA: Right Atrium
RHC: Right Heart Catheterization
RV: Right Ventricle
V_0_: unstressed RV volume
WSPH: World Symposium of Pulmonary Hypertension

## Author contributions

R.R. Vanderpool, conceived the idea for the study design and analyses detailed in this manuscript. S. Agarwal and R.R. Vanderpool contributed to the data collection and analysis in the study. R.R. Vanderpool performed the statistical analyses. All authors contributed to drafting and critical review of the manuscript. All authors approved the manuscript for submission.

## Acknowledgements

None

