## Supplemental Tables and Figures for "*RV IsoMax:* Development of a web-based single-beat analysis tool for the determination of RV-PA coupling"

### Supplemental Data

**Table S1.** Table of equations used to calculate single-beat RV function. Because RV volumes from cardiac magnetic resonance imaging were available, End-systolic elastance and arterial elastance were calculated using end-diastolic volume (EDV) and end-systolic volume (ESV). Abbreviations: RV: right ventricle, RVESP: end systolic pressure, CO: cardiac output, HR: Heart rate, Pmax: maximum isovolumetric pressure, and RVEF: RV ejection fraction

| Variables | Equation | Units |
|---|---|---|
| **Pulmonary Pressure/Afterload** | | |
| Ea: Arterial Elastance | (RVESP)/(EDV – ESV) | mmHg/ml |
| **Blood Flow** | | |
| CO: Cardiac Output | Thermodilution/Fick | L/min |
| SV: Stroke Volume | CO/HR*1000 and/or EDV-ESV | ml/beat |
| **RV Systolic/Global Function** | | |
| Ees: End-systolic Elastance | (Pmax- RVESP)/(EDV-ESV) | mmHg/ml |
| V0: Unstressed volume of the RV | [(Ees*ESV)-RVESP]/Ees | ml |
| Ees/Ea: RV-PA coupling | (Pmax/ESP) – 1 | ~ |
| $RVEF_{est}$: Pressure-only RVEF | 1-(ESP/Pmax) | % |

### Supplemental Figures

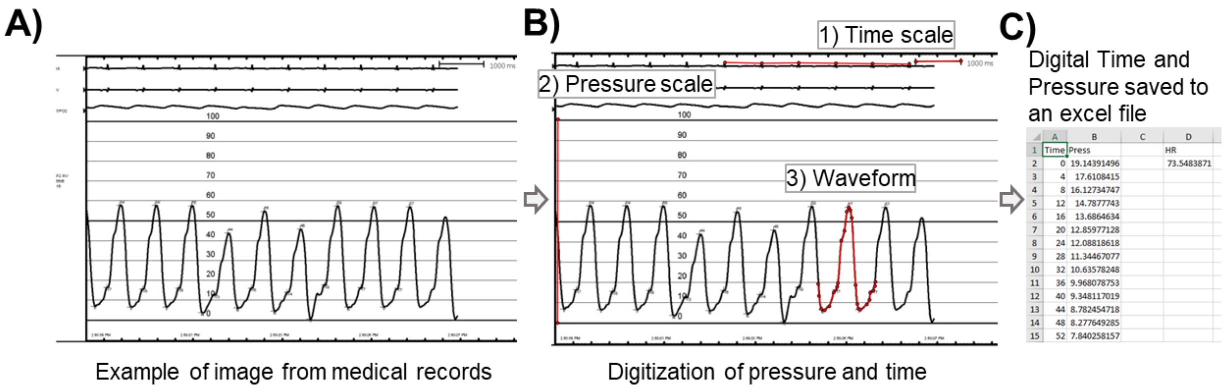


**Figure S1. Digital Conversion of RV Pressure Screen Captures. A)** Screen capture of the RV waveform in the medical records from a right heart catheterization. **B)** The custom Matlab digitization program allows the user to 1) set the time scale, 2) set the pressure scale and 3) select points along the pressure waveform to re-digitize the waveform. **C)** The digitized time and pressure and then saved to an excel file with time in milliseconds (ms), pressure in mmHg. Heart rate is calculated based on user selected points on electrocardiogram (ECG).

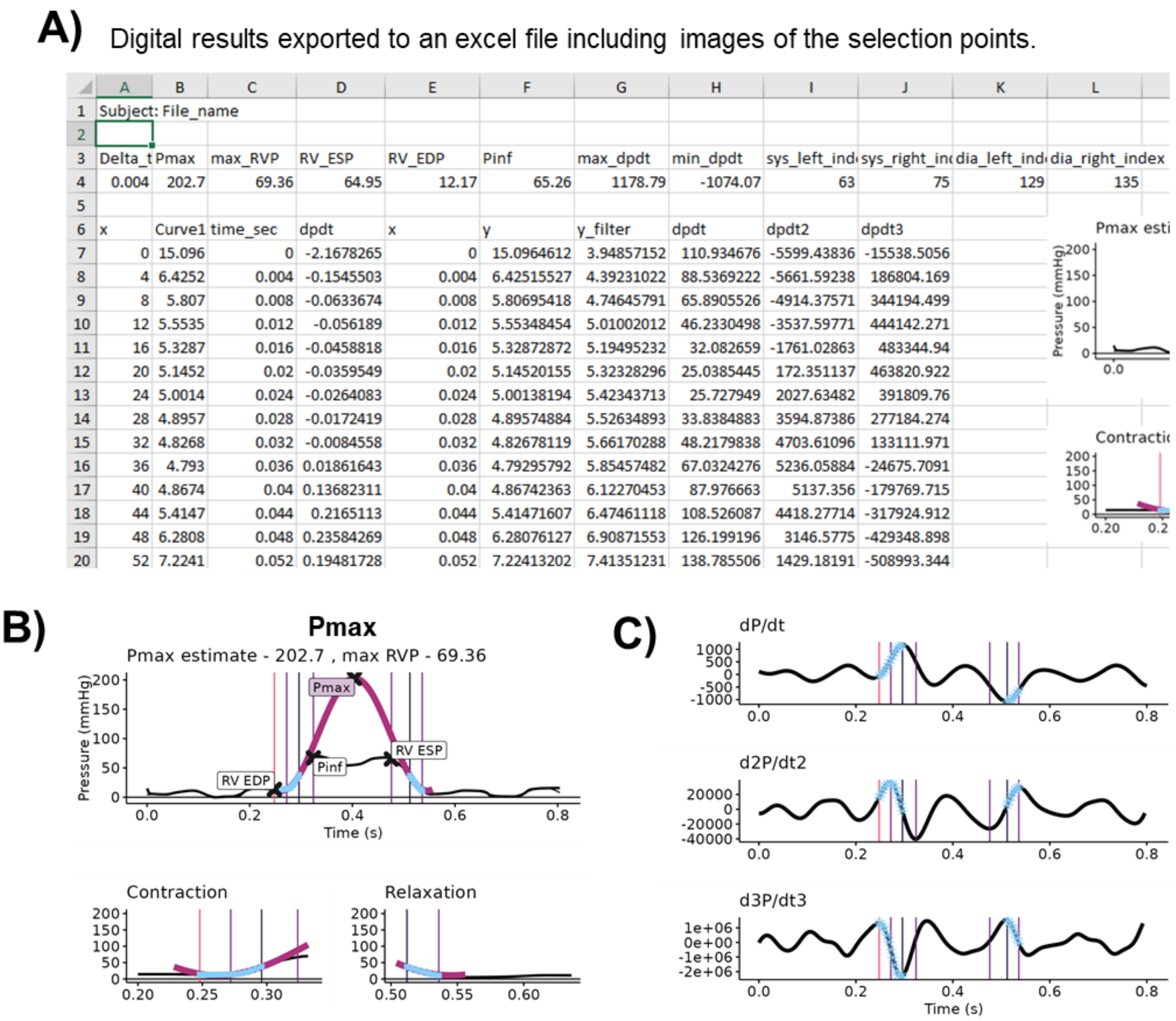

A) Digital results exported to an excel file including images of the selection points.

| | A | B | C | D | E | F | G | H | I | J | K | L |
|---|---|---|---|---|---|---|---|---|---|---|---|---|
| 1 | Subject: File_name | | | | | | | | | | | |
| 2 | | | | | | | | | | | | |
| 3 | Delta_t | Pmax | max_RVP | RV_ESP | RV_EDP | Pinf | max_dpdt | min_dpdt | sys_left_ind | sys_right_in | dia_left_ind | dia_right_index |
| 4 | 0.004 | 202.7 | 69.36 | 64.95 | 12.17 | 65.26 | 1178.79 | -1074.07 | 63 | 75 | 129 | 135 |
| 5 | | | | | | | | | | | | |
| 6 | x | Curve1 | time_sec | dpdt | x | y | y_filter | dpdt | dpdt2 | dpdt3 | | |
| 7 | 0 | 15.096 | 0 | -2.1678265 | 0 | 15.0964612 | 3.94857152 | 110.934676 | -5599.43836 | -15538.5056 | | |
| 8 | 4 | 6.4252 | 0.004 | -0.1545503 | 0.004 | 6.42515527 | 4.39231022 | 88.5369222 | -5661.59238 | 186804.169 | | |
| 9 | 8 | 5.807 | 0.008 | -0.0633674 | 0.008 | 5.80695418 | 4.74645791 | 65.8905526 | -4914.37571 | 344194.499 | | |
| 10 | 12 | 5.5535 | 0.012 | -0.056189 | 0.012 | 5.55348454 | 5.01002012 | 46.2330498 | -3537.59771 | 444142.271 | | |
| 11 | 16 | 5.3287 | 0.016 | -0.0458818 | 0.016 | 5.32872872 | 5.19495232 | 32.082659 | -1761.02863 | 483344.94 | | |
| 12 | 20 | 5.1452 | 0.02 | -0.0359549 | 0.02 | 5.14520155 | 5.32328296 | 25.0385445 | 172.351137 | 463820.922 | | |
| 13 | 24 | 5.0014 | 0.024 | -0.0264083 | 0.024 | 5.00138194 | 5.42343713 | 25.727949 | 2027.63482 | 391809.76 | | |
| 14 | 28 | 4.8957 | 0.028 | -0.0172419 | 0.028 | 4.89574884 | 5.52634893 | 33.8384883 | 3594.87386 | 277184.274 | | |
| 15 | 32 | 4.8268 | 0.032 | -0.0084558 | 0.032 | 4.82678119 | 5.66170288 | 48.2179838 | 4703.61096 | 133111.971 | | |
| 16 | 36 | 4.793 | 0.036 | 0.01861643 | 0.036 | 4.79295792 | 5.85457482 | 67.0324276 | 5236.05884 | -24675.7091 | | |
| 17 | 40 | 4.8674 | 0.04 | 0.13682311 | 0.04 | 4.86742363 | 6.12270453 | 87.976663 | 5137.356 | -179769.715 | | |
| 18 | 44 | 5.4147 | 0.044 | 0.2165113 | 0.044 | 5.41471607 | 6.47461118 | 108.526087 | 4418.27714 | -317924.912 | | |
| 19 | 48 | 6.2808 | 0.048 | 0.23584269 | 0.048 | 6.28076127 | 6.90871553 | 126.199196 | 3146.5775 | -429348.898 | | |
| 20 | 52 | 7.2241 | 0.052 | 0.19481728 | 0.052 | 7.22413202 | 7.41351231 | 138.785506 | 1429.18191 | -508993.344 | | |



Output of pressure and sinusoidal fit from RV IsoMax program/App

**Figure S2. Results saved in the exported Excel file in the *RV IsoMax* program. A)** The saved excel file includes the file name as the subject id. Program results are saved in Row 4 including Pmax and landmarks on the RV waveform. **B)** Example of the figure saved of the Sine fit (Purple curve) on the RV waveform including Pmax and landmark location. Fit regions are identified by light blue symbols. Vertical lines represent the time landmarks derived from pressure derivatives. From left to right: **Contraction:** max $d^3P/dt^3$, max $d^2P/dt^2$, max dP/dt, min $d^2P/dt^2$ and **Relaxation:** min $d^2P/dt^2$, min dP/dt, max $d^2P/dt^2$ **C)** 1st, 2nd and 3rd derivative of pressure that were used to identify fit regions. Abbreviations: delta_t: sampling time step, Pmax: isovolumetic max pressure, max RVP: maximum RV pressure, RV_ESP: RV end-systolic pressure, RV_EDP: RV end-diastolic pressure, Pinf: RV pressure at the 1$^{st}$ inflection during contraction, max_dP/dt: maximum dP/dt, min_dP/dt: minimum dP/dt. sys_left_index: index of the start of contraction region, sys_right_index: index used at the end of the contraction region, dia_left_index: index of the start of the relaxation region, dia_right_index: index of the end of the relaxation region.
